# Relationship between five Epigenetic Clocks, Telomere Length and Functional Capacity assessed in Older Adults: Cross-sectional and Longitudinal Analyses

**DOI:** 10.1101/2021.10.05.21264547

**Authors:** Valentin Max Vetter, Christian Humberto Kalies, Yasmine Sommerer, Dominik Spira, Johanna Drewelies, Vera Regitz-Zagrosek, Lars Bertram, Denis Gerstorf, Ilja Demuth

## Abstract

DNA methylation age acceleration (DNAmAA, derived from an epigenetic clock) and relative leukocyte telomere length (rLTL) are widely accepted biomarkers of aging. Nevertheless, it is still unclear which aspects of aging they represent best. Here we evaluated longitudinal associations between baseline rLTL and DNAmAA (estimated with 7-CpG clock) and functional assessments covering different domains of aging. Additionally, we made use of cross-sectional data on these assessments and examined their association with DNAmAA estimated by five different DNAm age measures.

Two-wave longitudinal data was available for 1,083 participants of the Berlin Aging Study II (BASE-II) who were re-examined on average 7.4 years after baseline as part of the GendAge study. Functional outcomes were assessed with Fried’s frailty score, Tinetti mobility test, falls in the past 12 months (yes/no), Finger-floor distance, Mini Mental State Examination (MMSE), Center for Epidemiologic Studies Depression Scale (CES-D), Activities of Daily Living (ADL), Instrumented ADL (IADL) and Mini Nutritional Assessment (MNA).

Overall, we found no evidence for an association between the molecular biomarkers measured at baseline, rLTL and DNAmAA (7-CpG clock), and functional assessments assessed at follow-up. Similarly, a cross-sectional analyses of follow-up data did also not show evidence for associations of the various DNAmAA measures (7-CpG clock, Horvath’s clock, Hannum’s clock PhenoAge, and GrimAge) with functional assessments.

In conclusion, neither rLTL nor 7-CpG DNAmAA were able to predict impairment in the analyzed assessments over a ∼7 year time-course. Similarly, DNAmAA as estimated by five epigenetic clocks was not a good cross-sectional marker of health deterioration either.

## Introduction

The identification of factors indicating healthy aging is an intensively studied aspect of gerontological research and has gained importance over the past decades due to demographic changes [1]. A well-studied biomarker is the length of telomeres, the protective end structures of linear chromosomes, which shorten with every cycle of DNA replication (reviewed in [2]). The comparatively new and lately intensively studied epigenetic age estimates, DNA methylation age (DNAm age) and particularly its deviation from chronological age, DNAm age acceleration (DNAmAA), were reported to be promising biomarkers as well [3, 4]. Several versions of epigenetic clocks are available that differ predominantly in the location and number of CpG sites included [5-8] as well as the method that is used to obtain information on DNA methylation. Additionally, epigenetic clocks seem to differentiate in aspects of aging they represent best [3, 9, 10].

Both biomarkers, rLTL and DNAmAA, are well evaluated with regard to mortality (reviewed for rLTL [11] and epigenetic clocks [12]) and cross-sectional associations with various age associated phenotypes and diseases are reported as well (reviewed for rLTL [11] and epigenetic clocks [4]). However, rLTL and DNAm age seem to be independent from each other since neither our group [13, 14] nor others [15] observed any noteworthy associations between both biomarkers.

Although the relationship of these markers with mortality and age-associated diseases is well documented, studies that systematically analyze their association with clinical assessments of cognitive and physical capacity are limited [9, 10]. We have previously reported on a cross-sectional analysis of Fried’s frailty score, assessments for physical and mental health, cognitive capacity with an epigenetic clock that is based on seven CpG sites and was trained in 1,895 participants of the Berlin Aging Study II (BASE-II) [13]. Overall, we did not find evidence for cross-sectional associations of the biomarkers with functional status in this previous analysis at baseline [16]. This could be explained, at least in part, by the fact that BASE-II participants were predominantly healthy at baseline, resulting in a low proportion of participants reporting impairment.

These previous results, as well as most other studies on this topic, used a cross-sectional analysis scheme. However, several authors have emphasized the necessity of longitudinal analyses [3, 17]. Furthermore, the potential utility of DNAmAA as longitudinal biomarker was shown only recently for Horvath’s and Hannum’s clocks in a meta-analysis of five longitudinal cohorts [18]. Nevertheless, the literature on longitudinal studies investigating the relationship between DNAmAA and age-related outcomes over time is currently very limited [9, 19]. In this study, we present longitudinal data on the 7-CpG epigenetic clock and examine the longitudinal relationship between rLTL and the 7-CpG DNAmAA assessed at baseline and several variables of mental and physical health, including the Fried’s frailty phenotype, that were assessed after up to ten years of follow-up [20]. Subsequently, we analyzed the cross-sectional relationship between functional assessments and DNAmAA as calculated from five different DNAm age estimates at follow-up.

Aging occurs differently in women and men [21] and sex-differences in epigenetic aging have been repeatedly reported [22, 23]. Therefore, all analyses were performed in the whole dataset as well as within sex-stratified subgroups.

## Methods

### Berlin Aging Study II (BASE-II) and GendAge Study

The Berlin Aging Study II (BASE-II) is a multi-institutional and multi-disciplinary study that aims to identify factors associated with “healthy” vs. “unhealthy” aging. A convenience sample of 1,671 older residents aged 60-85 years (plus a younger control group, n= 500, not considered here) of the greater metropolitan area of Berlin, Germany, was assessed at baseline in the medical part of the study [24, 25]. Participants of the older age group were medically re-assessed on average 7.4 years later as part of the GendAge study [20]. Follow-up data were available for 1,083 individuals. Seventeen additional participants were not medically assessed at baseline but were available for the cross-sectional analysis at follow-up (n=1,100). Availability of variables for baseline and follow-up examination are illustrated in Supplementary Figure 1.

All participants gave written informed consent. The medical assessments at baseline and follow-up were conducted in accordance with the Declaration of Helsinki and approved by the Ethics Committee of the Charité – Universitätsmedizin Berlin (approval numbers EA2/029/09 and EA2/144/16) and were registered in the German Clinical Trials Registry as DRKS00009277 and DRKS00016157.

### Relative Leukocyte Telomere Length (rLTL)

A modified version of the protocol by Cawthon was used to assess rLTL in participants at baseline. Briefly, triplicates of single copy gene PCR (36B4) and telomere PCR were performed in 384-well plate format with each available leukocyte DNA sample. Leukocyte DNA of 10 randomly selected BASE-II participants was used as reference sample on every plate. Telomer length was calculated and Pfaffl correction considered. For detailed information on the measurement of rLTL in BASE-II, please refer to Meyer et al. [26].

### Methylation-sensitive Singe Nucleotide Primer Extension (MS-SNUPE)

DNAm age was estimated with the 7-CpG clock measured at baseline and follow-up from DNAm data obtained using a modified version of the methylation-sensitive single nucleotide primer extension (MS-SNuPE) protocol for methylation age [27, 28], which we described in detail elsewhere (for baseline [13] and follow-up [16]).

Briefly, 1000 ng of leukocyte DNA were bisulfite converted and subsequently amplified by multiplex PCR. A single nucleotide primer extension (SNuPE) was performed on the PCR products. Subsequently, the methylation fraction of sites of interest was measured from 6 μg of SNuPE products by a “3730 DNA Analyzer” (Applied Biosystems, HITACHI). Finally, DNAm age was calculated by the methylation fraction of seven cytosine-phosphate-guanine (CpG) sites. The measurement of baseline and follow-up samples were conducted in the same laboratory and by the same protocol which only differed in the amount of DNA that was used for bisulfite conversion and the amount of SNuPE products that was measured in the last step.

### Infinium MethylationEPIC array

We also determined genome-wide DNAm profiles using the “Infinium MethylationEPIC” array (Illumina, Inc, USA). These data were only available for DNA samples obtained during follow-up (Supplementary Figure 1). Data pre-processing and QC was performed in R (v3.6.1 [29]) using the *bigmelon* package. In brief, we excluded CpG probes that had ≥1% samples with a detection p-value of 0.05 or a bead count below three in >5% of samples. Efficiency of bisulfite conversion was assessed with *bscon* (samples with values <80% were excluded). After exclusion of outliers, samples were reloaded and normalized with the *dasen* function. Changes in beta values due to normalization were assessed with the *qual* function and samples with a deviation |≥0.1| in beta values after normalization were removed. After removal of excluded CpG probes, the new dataset was reloaded and normalized again and subjected to DNAm age estimation (see below).

### DNA Methylation Age (DNAm age) and DNAm Age Acceleration (DNAmAA)

DNAm age was estimated from five different versions of epigenetic clocks. The 7-CpG clock formula was trained on participants of the BASE-II cohort at baseline [13]. Although it was specifically developed for SNuPE methylation data, a newly available adjustment formula allows it to be calculated from Illumina methylation data as well [30].

Horvath’s clock, Hannum’s clock, PhenoAge and GrimAge were calculated according to the manual on Steve Horvath’s website (https://horvath.genetics.ucla.edu/html/dnamage/). Data on one CpG site of PhenoAge and on seven CpG sites of Hannum’s clock were not available for DNAm age calculation in the BASE-II dataset.

DNAm age acceleration (DNAmAA) was calculated as “intrinsic epigenetic age acceleration (IEAA)” [31] as residuals of a linear regression analysis of DNAm age on chronological age and cell counts (neutrophils, monocytes, lymphocytes, eosinophils). Leukocyte cell type composition was measured by flow cytometry in an accredited standard laboratory.

### Frailty score and functional assessments

To measure frailty in this study we used a slightly adapted version of Fried’s frailty score [32] which considers unintentional weight loss, self-reported exhaustion, weakness (grip strength), slow walking speed and low physical activity resulting in a score between 0 (no impairment) and 5 (impairment in all assessed domains) [33]. Cut-off for impairment was a frailty score higher than 0 [34].

Tinetti mobility test, falls in past 12 months, finger-floor distance, Mini-Mental State Examination (MMSE), Center for Epidemiologic Studies Depression Scale (CES-D), Activities of Daily Living (ADL), Instrumental ADL (IADL) and Mini Nutritional Assessment (MNA) were analyzed in this study. For logistic regression analyses and a more structured display of the data, assessment scores were dichotomized according to evaluated cut-off values (for a detailed display on range of scores and established cut-off values, please refer to Supplementary Table 1 of ref. [16]). As proposed by Lawton and Brody [35] a sex stratified assessment of IADL was conducted (food preparation, housekeeping, laundering were not included in the questionnaire for men). We used two or more dependencies as cut-off value to define impairment for IADL [36]. All tests were conducted by trained study personnel.

### Covariates

We included the following covariates in all regression analyses based on previous evidence suggesting a potential effect of these variables on both DNAm patterns and the studied outcomes [3]: 1. sex (male/female). 2. Alcohol consumption; this was assessed in g/d at baseline (via food frequency questionnaire [37]) and as “yes/no”-question at follow-up. 3). Smoking behavior; documented in packyears in one-to-one interviews. 4. Height and weight; measured using the electronic measuring station “seca 763” (SECA, Germany) and used to calculate the body mass index (BMI). 5. Genetic ancestry; quantified via principal component analysis on genome-wide SNP genotyping data [38]; here, the first four principal components were included to account for subtle differences in population substructure. Since chronological age was not correlated with any of the DNAmAA measures (Pearson’s r <0.1, data not shown) it was not included as covariate.

The morbidity index was computed as a modified version [39] of the Charlson index [40]. All longitudinal regression models were adjusted for covariates that were assessed at baseline and the cross-sectional analysis included covariates that were measured at follow-up.

### Statistical analysis

Statistical analyses were performed with R 3.6.2 [29]. Differences in proportions of impairment between baseline and follow-up were assessed by McNemar’s Test for paired datasets. Regression analyses were calculated with the “glm” function of R’s “stats” library. Figures were created with the “ggplot2” package [41] and its extension “GGally” and “UpSetR” [42]. We defined a p-value below 0.05 as statistically significant. Because no statistically significant results were found, we did not elaborate further on multiple testing. An available case analysis (pairwise deletion) was employed for all statistical analyses. Therefore, participants were excluded only from an analysis if they were missing one or more variables that were relevant to the performed analyses. We indicated the number of observations for each individual analysis.

## Results

### Study population and results of functional assessments in follow-up examinations

One thousand eighty-three participants of the Berlin Aging Study II (BASE-II) were medically re-examined on average 7.4 years (SD = 1.5, range: 3.9 years – 10.4 years) after baseline examination. The mean age at follow-up was 75.6 years (SD = 3.77, age range: 64.9 - 94.1 years) and men and women were almost equally distributed (52% female). Participant characteristics at baseline and follow-up are shown in Table 1 and Supplementary Table 1.

**Table 1:** Descriptive statistics of participants at baseline (T0) and follow-up (T1).

|  | Baseline (2009-2014) |  |  |  |  |  | Follow-up (2018-2020) |  |  |  |  |  |
| --- | --- | --- | --- | --- | --- | --- | --- | --- | --- | --- | --- | --- |
|  | n | % | Mean | SD | Min | Max | n | % | Mean | SD | Min | Max |
| Chronological Age | 1083 |  | 68.27 | 3.48 | 60.16 | 84.63 | 1100 |  | 75.60 | 3.77 | 64.91 | 94.07 |
| Sex (female) | 1083 | 51.99 |  |  |  |  | 1100 | 52.09 |  |  |  |  |
| 7-CpG DNAm Age | 971 |  | 65.78 | 7.45 | 40.25 | 91.81 | 1092 |  | 71.21 | 7.17 | 41.65 | 109.18 |
| 7- CpG DNAmAA | 922 |  | -0.10 | 6.82 | -22.93 | 22.33 | 1071 |  | 0.03 | 6.42 | -24.87 | 34.42 |
| rLTL | 927 |  | 1.147 | 0.230 | 0.167 | 1.921 |  |  |  |  |  |  |
| Smoking (packyears) | 933 |  | 9.89 | 17.09 | 0.00 | 148.00 | 1019 |  | 9.79 | 17.61 | 0.00 | 150.00 |
| Morbidity Index | 986 |  | 0.97 | 1.20 | 0.00 | 7.00 | 954 |  | 1.39 | 1.54 | 0.00 | 9.00 |
| BMI | 957 |  | 26.65 | 4.07 | 17.69 | 47.68 | 1098 |  | 26.97 | 4.25 | 17.17 | 49.68 |
| Alcohol Intake (g/d) | 957 |  | 15.05 | 17.55 | 0.39 | 120.78 |  |  |  |  |  |  |
| Alcohol Intake (yes) |  |  |  |  |  |  | 1097 | 83.14 |  |  |  |  |

Seventeen BASE-II participants were not medically assessed at baseline but examined at follow-up and are used in all cross-sectional analyses of follow-up data.
Note: DNAm: deoxyribonucleic acid methylation; rLTL: relative leukocyte telomere length; BMI: body mass index.

The highest proportion of impairment at follow-up among the assessments analyzed here was detected in the finger-floor distance test (62.2% were not able to reach the ground with their fingertips, Table 2) which additionally was, together with Fried’s frailty score, included in the most frequent impairment-combination (Figure 1 A). Both assessments have the highest proportions of participants that show a singular impairment: 175 participants had their only impairment in the finger-floor distance test and 85 participants were only impaired with respect to Fried’s frailty score (Figure 1 A, Supplementary Table 2). The lowest proportions of isolated impairments among all participants were found in the Tinetti Mobility Test (0.3%), MMSE (0.1%) and ADL (0.3%), and none of the participants had a singular impairment in the IADL. Overall, only 26.9% of participants showed impairment in three or more assessments at follow-up (Figure 1 C). Sex-stratified analyses are shown in Figure 1 D and E and Supplementary Table 3. In both baseline and follow-up, more than two thirds of participants without any impairment were female, while men and women were more equally distributed in groups with one or more impairments (Figure 1 D and E, Supplementary Table 3).

**Table 2:** Descriptive statistics of assessments at baseline (T0) and follow-up (T1).

| Assessment | n | Baseline (2009-2014) |  |  | Follow-up (2018-2020) |  |  | p-value |  |
| --- | --- | --- | --- | --- | --- | --- | --- | --- | --- |
|  |  | mean | SD | impaired (%) | mean | SD | impaired (%) |  |  |
| Frailty score | 905 | 0.34 | 0.59 | 28.51 | 0.77 | 0.88 | 53.48 | <0.001 | *** |
| Tinetti mobility test | 1010 | 27.66 | 1.48 | 1.39 | 26.58 | 2.63 | 7.82 | <0.001 | *** |
| Falls in past 12 months | 833 |  |  | 34.57 |  |  | 27.25 | <0.001 | *** |
| Finger-floor distance | 1062 | 9.66 | 11.75 | 57.63 | 10.52 | 12.63 | 62.24 | <0.001 | *** |
| MMSE | 1016 | 28.59 | 1.32 | 1.28 | 28.57 | 1.53 | 1.87 | 0.377 |  |
| CES-D | 1043 | 13.93 | 3.5 | 27.52 | 13.57 | 3.67 | 23.97 | 0.040 | * |
| ADL | 1071 | 99 | 3.74 | 2.15 | 98.71 | 4.09 | 4.58 | <0.001 | *** |
| IADL | 1070 |  |  | 0.19 |  |  | 0.65 | 0.182 |  |
| MNA | 1021 | 27.38 | 1.75 | 2.84 | 26.62 | 2.15 | 10.97 | <0.001 | *** |

McNemar's test was used to test for difference of proportion between baseline and follow-up. Only participants whose data were available for both baseline and follow-up were included in this table. Due to the scale differences between the genders, only the dichotomized values are displayed for

**Figure 1:**
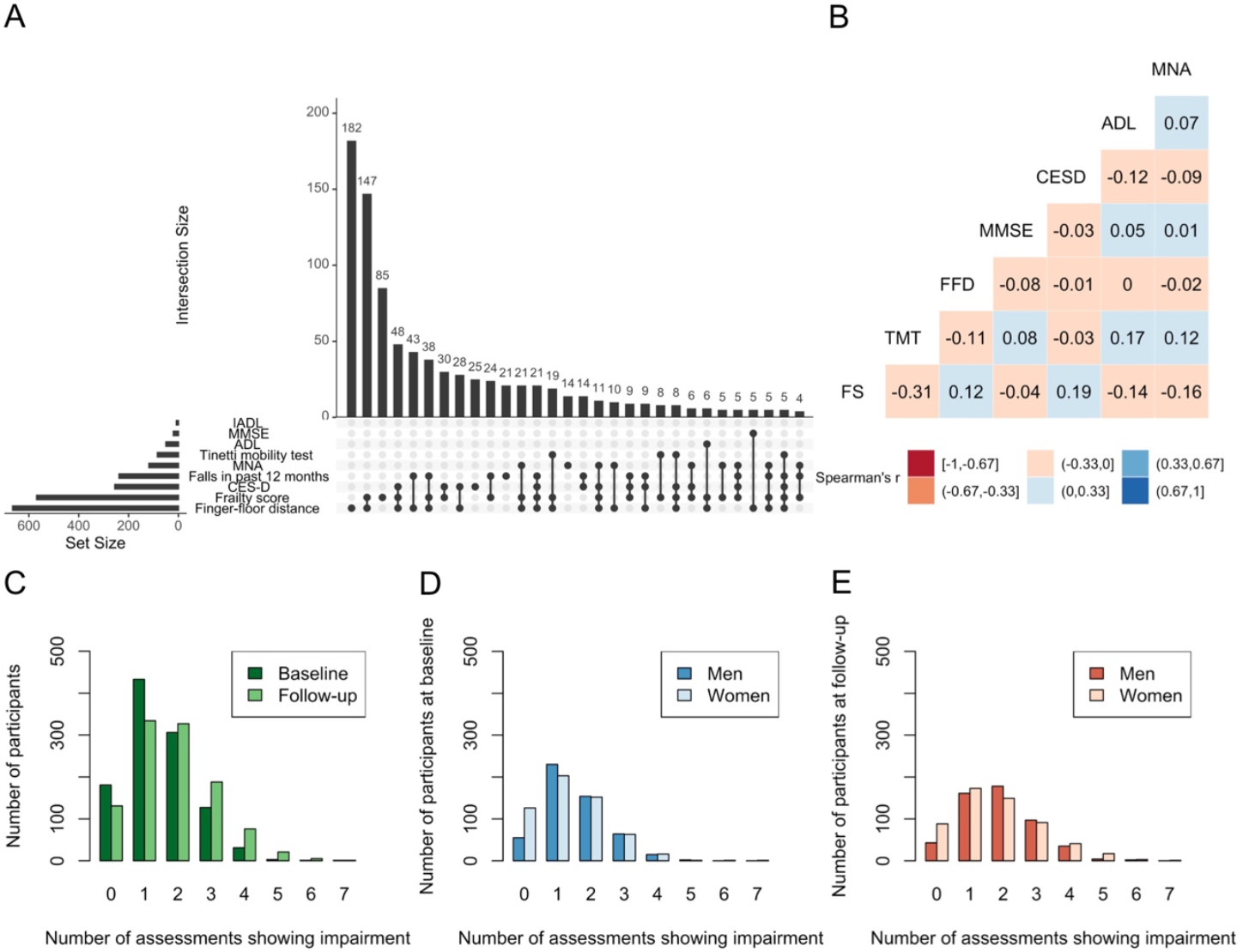
Frequency of impairment and correlation between assessments/frailty score. A: Upset plot of the thirty most frequent combinations of impairment at follow-up. The most frequent combination was found for the frailty score and finger-floor distance. B: Heatmap of Spearman’s correlation between assessment scores at follow-up. C-E: Barplots of number of participants per number of assessments reflecting impairment stratified by timepoint of measurement (C) and by sex at (D) baseline and (E) follow-up. Note: FS: Frailty Score, TMT: Tinetti mobility test, FFD: Finger-floor distance, MMSE: Mini-Mental State Examination, CESD: Center for Epidemiologic Studies Depression Scale, ADL: Activities of Daily Living, IADL: Instrumented ADL, MNA: Mini Nutritional Assessment.

### Longitudinal data on the 7-CpG clock

Seven-CpG DNAm age was 65.8 years (SD = 7.5 years) at baseline and 71.2 years (SD = 7.2 years) at follow-up. Pearson correlation between chronological age and 7-CpG DNAm age was r=0.31 at baseline and r=0.29 at follow-up in the subgroup of older BASE-II participants studied here. The average slope of the relative change of DNAm age to chronological age between the two assessed timepoints was 0.75 (SD = 0.64, range: -4.8 to 5.4, n=965). Therefore, in terms of epigenetic age participants aged on average 0.75 years for every additional chronological year. Slower epigenetic aging compared to elapsed time, especially in older cohorts, was reported before [43]. The test-retest correlation (rank order stability) was high (Pearson’s r=0.81, n=965).

For a better understanding of the longitudinal progression, we divided the dataset into groups. Participants with negative slope are biologically younger at follow-up than at baseline and are therefore classified as “rejuvenated”. Participants with a slope of 1 aged at the same pace chronologically and biologically. To account for random error and measurement error in DNAm age, we defined a slope of 0.76 to 1.25 as “adequately aged”. Participants with a slope > 1.25 were classified as “prematurely aged” and a slope between 0 and 0.75 was indicative for “slow aging” (Figure 2 A). Please note that the limits for “adequate aging” are somewhat arbitrary but were chosen for a more structured display of the longitudinal data.

**Figure 2:**
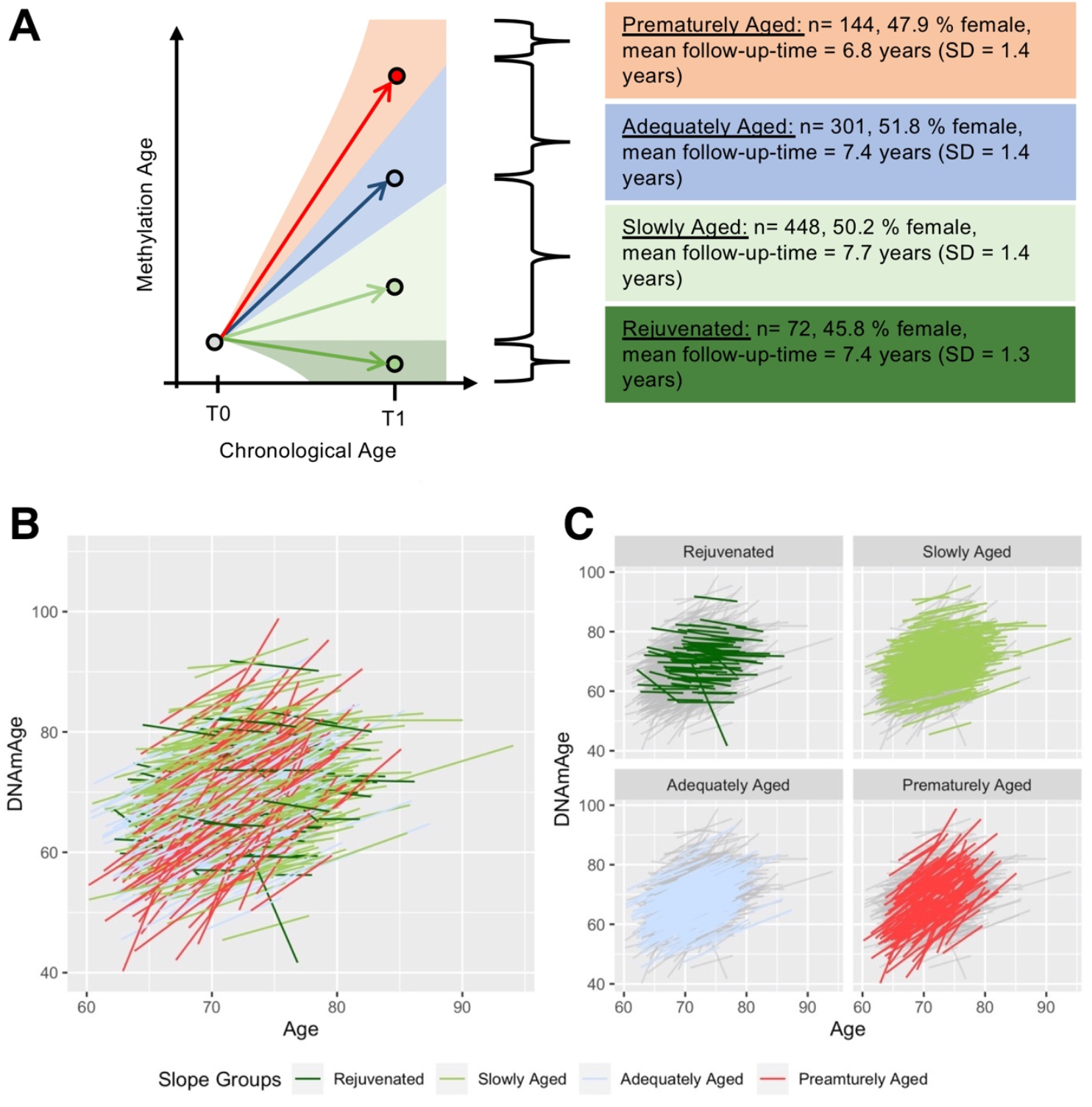
**Possible trajectories of an individual’s DNAm age between baseline (T0) and follow-up (T1) examination (A) and Spaghetti Plots of the longitudinal DNAm age data of older participants of the BASE-II/GendAge cohort (B, C).** For a more structured display of the DNAm age trajectories we identified four possible groups that were defined based on the slope: prematurely aged (> 1.25), adequately aged (0.76-1.25), slowly aged (0-0.75) and rejuvenated (< 0). Please note that these cut-off values are somewhat arbitrary and were chosen to allow a form of stratification. Besides the schematic display of possible trajectories (A), the actually observed DNAm age trajectories are shown as spaghetti plots with all participants (B) as well as stratified by groups (C).

Using these criteria, we found 14.9% of participants to be “prematurely aged” at follow-up. Only one third (31.2%) aged adequately according to our definition. Almost half of the analyzed participants (46.4%) were found to have aged slowly and 7.5% were biologically younger at follow-up compared to baseline (Figure 2).

The slope of change did not correlate with time between measurement points (Pearson’s r=-0.16).

### Change in functional assessments over up to 10 years of follow-up

The fraction of participants with test results below the established cut-off values (and thus categorized as *impaired*; for more details please see Supplementary Table 1 in ref. [16]) increased significantly between baseline and follow-up examinations in almost all functional assessments (Table 2). The proportion of participants that were pre-frail or frail as assessed by Fried’s frailty score increased from 28.5% to 53.5% between baseline and follow-up (p<0.001), which is the highest difference in assessment results between baseline and follow-up in this study. The second highest difference was found in the Mini Nutritional Assessment results (impairment in 2.8% vs. 11.0%, p<0.001, Table 2). Interestingly, participants reported fewer symptoms of depression as assessed by the CES-D (below cut-off: 27.5% vs. 24.0%, p=0.04) and falls within the past 12 months (reported ≥1 fall: 34.6% vs. 27.3%, p<0.001) at follow-up when compared to baseline. No change over the observation period was detected in the average results of a global screening test on cognitive function, the MMSE (below cut-off: 1.28% vs. 1.87%, p=0.377) and the IADL (below cut-off: 0.2% vs 0.7%, p=0.18). Due to the low variance of IADL, it was excluded from all further analyses.

The average number of assessments per person showing impairment increased by 0.38 (Supplementary Table 3). While 16.7% of the participants were not impaired at baseline, only 12.1% were without any impairment at follow-up (Figure 1 C, Supplementary Table 3).

Correlation between the continuous assessment scores assessed at follow-up was weak (r≤|0.31|, Figure 1 B). The strongest correlation (r=-0.31) was found between Fried’s frailty score (higher scores indicating higher degree of impairment) and the Tinetti mobility test (lower scores indicating higher degree of impairment).

### Longitudinal analyses: 7-CpG DNAmAA and rLTL are not associated with impairment in functional assessments after up to ten years of follow-up

Baseline data on the biomarkers of aging, rLTL and 7-CpG DNAmAA, was available for 927 and 922 of the participants who were examined at follow-up. We performed multiple linear regression analyses of each assessment score (assessed at follow-up) on rLTL and 7-CpG DNAmAA (assessed at baseline) and adjusted for genetic PCs, sex, alcohol consumption, smoking (packyears), morbidity index and body mass index (BMI), all assessed at baseline. The analyses revealed that neither rLTL nor 7-CpG DNAmAA were associated with any of the future functional outcomes (Table 3). Sex-stratified subgroup analyses revealed a statistically significant but minimal association between ADL and 7-CpG DNAmAA in men (*multiple linear regression*, Supplementary Table 5) that were too small to be of clinical relevance (difference per year of age acceleration below 0.1% of the respective scale range).

**Table 3:** Longitudinal analyses: Individual multiple linear regression analyses of functional assessments at follow-up on baseline rLTL and 7-CpG DNAmAA.

| Assessments | $\beta$ | SE | p-value | n |
| --- | --- | --- | --- | --- |
| rLTL |  |  |  |  |
| Frailty score | 0.08 | 0.14 | 0.54 | 749 |
| Tinetti mobility test | -0.02 | 0.39 | 0.95 | 743 |
| Finger-floor distance | -0.99 | 1.97 | 0.62 | 749 |
| MMSE | 0.05 | 0.26 | 0.85 | 750 |
| CES-D | 0.75 | 0.59 | 0.21 | 751 |
| ADL | 0.00 | 0.69 | 0.99 | 754 |
| MNA | -0.34 | 0.36 | 0.34 | 738 |
| 7-CpG DNAm Age Acceleration |  |  |  |  |
| Frailty score | 0.00 | 0.00 | 0.67 | 713 |
| Tinetti mobility test | -0.01 | 0.01 | 0.29 | 708 |
| Finger-floor distance | -0.04 | 0.07 | 0.60 | 714 |
| MMSE | 0.00 | 0.01 | 0.63 | 714 |
| CES-D | -0.02 | 0.02 | 0.38 | 715 |
| ADL | -0.04 | 0.02 | 0.14 | 718 |
| MNA | 0.00 | 0.01 | 0.98 | 703 |

Individual multiple linear regression analyses were adjusted for genetic PCs, sex, alcohol consumption, smoking behavior, morbidity index and body mass index. Biomarkers of aging and all covariates were measured at baseline. Assessments were measure at follow-up.
Note: rLTL: relative leukocyte telomere length, MMSE: Mini-Mental State Examination, CES-D: Center for Epidemiologic Studies Depression Scale, ADL: Activities of Daily Living, MNA: Mini Nutritional Assessment.

To analyze whether future impairment defined by established cut-off values of each assessment rather than continuous decline in ability, was associated with rLTL/7-CpG DNAmAA, individual logistic regression analyses of dichotomized assessment scores were executed. Similar to the linear regression analyses, no statistically significant associations were found (Supplementary Table 4).

Linear regression analyses of change in assessment scores (between baseline examination and follow-up) on 7-CpG DNAmAA, assessment score at baseline, chronological age, sex, alcohol (yes/no), smoking (packyears), morbidity index and body mass index showed no convincing associations either (data not shown).

### Cross-sectional analyses: DNAmAA derived from five different epigenetic clocks were not associated with physical and cognitive capacity at follow-up in BASE-II

We found no cross-sectional association in multiple linear regression analyses of each assessment score on DNAmAA of 7-CpG clock, Horvath’s clock, Hannum’s clock, PhenoAge or GrimAge after adjustment for covariates (Table 4). The only exception was a statistically significant but very weak association between GrimAge IEAA and Fried’s Frailty score (β = 0.02, SE = 0.01, p=0.03).

**Table 4:** Cross-sectional analyses: Individual multiple linear regression analyses of functional assessments on DNAmAA of five different epigenetic clocks and covariates (all measured at follow-up).

|  | Frailty score |  |  |  | Tinetti Mobility Test |  |  |  | Finger-floor distance |  |  |  | MMSE |  |  |  |
| --- | --- | --- | --- | --- | --- | --- | --- | --- | --- | --- | --- | --- | --- | --- | --- | --- |
| | $\beta$ | SE | p | n | $\beta$ | SE | p | n | $\beta$ | SE | p | n | $\beta$ | SE | p | n |
| 7-CpG IEAA | 0.00 | 0.00 | 0.82 | 789 | 0.01 | 0.01 | 0.62 | 777 | -0.02 | 0.07 | 0.72 | 788 | 0.00 | 0.01 | 0.89 | 786 |
| Horvath's IEAA | 0.00 | 0.01 | 0.62 | 787 | -0.01 | 0.02 | 0.78 | 775 | -0.13 | 0.11 | 0.20 | 786 | 0.00 | 0.01 | 0.87 | 783 |
| Hannum's IEAA | 0.00 | 0.01 | 0.73 | 787 | -0.01 | 0.02 | 0.52 | 775 | -0.07 | 0.11 | 0.52 | 786 | -0.02 | 0.01 | 0.25 | 783 |
| PhenoAge IEAA | 0.00 | 0.01 | 0.47 | 787 | 0.01 | 0.02 | 0.73 | 775 | -0.06 | 0.08 | 0.46 | 786 | 0.01 | 0.01 | 0.32 | 783 |
| GrimAge IEAA | 0.02 | 0.01 | 0.03 * | 787 | -0.03 | 0.03 | 0.37 | 775 | -0.06 | 0.14 | 0.65 | 786 | -0.01 | 0.02 | 0.51 | 783 |
|  | CES-D |  |  |  | ADL |  |  |  | MNA |  |  |  |  |  |  |  |
| | $\beta$ | SE | p | n | $\beta$ | SE | p | n | $\beta$ | SE | p | n | | | | |
| 7-CpG IEAA | 0.00 | 0.02 | 0.92 | 791 | -0.02 | 0.02 | 0.47 | 791 | -0.01 | 0.01 | 0.38 | 774 |  |  |  |  |
| Horvath's IEAA | 0.01 | 0.03 | 0.75 | 789 | -0.02 | 0.04 | 0.65 | 789 | -0.02 | 0.02 | 0.33 | 772 |  |  |  |  |
| Hannum's IEAA | -0.02 | 0.03 | 0.50 | 789 | -0.02 | 0.04 | 0.58 | 789 | -0.01 | 0.02 | 0.63 | 772 |  |  |  |  |
| PhenoAge IEAA | -0.04 | 0.02 | 0.12 | 789 | -0.01 | 0.03 | 0.68 | 789 | -0.01 | 0.01 | 0.38 | 772 |  |  |  |  |
| GrimAge IEAA | 0.01 | 0.04 | 0.82 | 789 | -0.07 | 0.05 | 0.18 | 789 | -0.03 | 0.03 | 0.19 | 772 |  |  |  |  |

All regression analyses were adjusted for genetic PCs, sex, alcohol consumption, smoking behavior, morbidity index and body mass index.
Assessment, DNAmAA and all covariates were measured at follow-up.
Note: IEAA: Intrinsic Epigenetic Age Acceleration, MMSE: Mini-Mental State Examination, CES-D: Center for Epidemiologic Studies Depression Scale, ADL: Activities of Daily Living, MNA: Mini Nutritional Assessment.

Sex-stratified analyses are displayed in Supplementary Table 6 and 7. The only statistically significant but weak association was found between Horvath’s DNAmAA and ADL in the female subgroup.

## Discussion

In this study, we made use of two-wave longitudinal data on eight markers of physical and mental health from older adults of the BASE-II cohort. We evaluated whether two biomarkers of aging, rLTL and DNAmAA assessed at baseline, were associated with future impairment in the assessments after up to 10.5 years (mean 7.4 years) follow-up. Subsequently, we repeated the previously reported cross-sectional analysis between functional assessments and 7-CpG clock with data from the follow-up examination [16] and now included DNAmAA derived from four other DNAm age estimates (Horvath’s clock, Hannum’s clock, PhenoAge and GrimAge, only available for follow-up examination).

Overall, the longitudinal analyses revealed no consistent associations between rLTL/7-CpG DNAmAA and functional assessments. We note that the body of literature on the potential relationship between the molecular biomarkers investigated here and longitudinal functional assessments is very limited. Cognitive decline measured by a modified version of the MMSE was reported to be associated with short leukocyte telomere length (LTL) [44], while longitudinal associations between LTL and MMSE [45] or lifetime cognitive change [46] were not reported. Marioni and colleagues [19] examined longitudinal data on Horvath DNAmAA and grip strength, walking speed and lung function but also failed to observe a significant association. Further, Maddock et al. analyzed the relationship between several measures of cognitive (episodic memory and mental speed) and physical (grip strength, chair rise speed and lung function) health and Horvath’s clock, Hannum’s clock, PhenoAge and GrimAge (n=973, age at baseline: 53 years, follow-up time: 16 years). After adjustment for cell composition, none of the first-generation clocks (trained to predict chronological age) was associated with future outcomes of physical/cognitive assessments but participants with higher PhenoAge or GrimAge showed worse performances in conducted tests [9]. Vershoor and colleagues found DNAmAA determined from Hannum’s clock and GrimAge to be associated with worse results in a 76-item frailty index [47]. Nevertheless, the comparability between the individual epigenetic clock constructs remains limited and the differences between previous and our findings could be the result of different epigenetic clocks used or differences in the frailty measures.

Our cross-sectional analyses of the BASE-II follow-up data revealed no association between analyzed assessments and the 7-CpG clock, Horvath’s clock, Hannum’s clock or PhenoAge. The only exception was a weak but nominally significant association between frailty and GrimAge. The size of the association means that GrimAge acceleration would need to be increased by more than 50 years to be associated with an increase by one point on the five-point Fried’s Frailty score. Future work needs to follow-up on this specific result. In the literature, a discrepancy in cross-sectional findings between first generation clocks (e.g. 7-CpG, Horvath, Hannum) and second generation clocks (PhenoAge, GrimAge) was reported before [9, 10, 47](overview in Supplementary Table 8). These previous studies reported a more distinct difference than observed here. Our observed lack of cross-sectional associations between the various mental and physical assessments and DNAm age estimated from first-generation clocks is in line with findings reported by Gale and colleagues [48]. These authors examined 791 70-year-olds (50.3% female) and reported significant associations between Fried’s frailty score and Hannum’s extrinsic epigenetic age acceleration (EEAA), but not with Horvath’s intrinsic epigenetic age acceleration (IEAA). Likewise, Kim and colleagues found a weak correlation between Horvath’s DNAm age and a 34-items Frailty Index (r=0.2, p<0.05) that did not remain significant after adjustment for chronological age [49]. In contrast, Breitling and colleagues reported a significant association between the same 34-item Frailty index and DNAmAA (Horvath’s clock) in a large cohort of 50 to 75 years old participants (n=1,820, 52% female). In this study, the frailty index increased by 1 deficit for every additional 12 years in methylation age acceleration [15]. Maddock et al. found no significant associations between Horvath’s or Hannum’s clock and physical (grip strength, chair rise speed) or mental performance (episodic memory and mental speed). After adjustment for covariates, associations between higher PhenoAge acceleration and chair rise speed and between GrimAge acceleration and cognitive capacity measures were statistically significant [9]. Similar to the results reported in this study, McCrory et al. found no association between several markers for cognitive and physical health and Horvath’s clock, Hannum’s clock or PhenoAge after covariate adjustment. GrimAge, however, was associated with walking speed and Fried’s frailty phenotype (after adjustment for covariate) [10]. In line with the results reported by Maddock and McCrory, Verschoor and colleagues reported only recently on a significant association between a 76-item frailty index and GrimAge acceleration that was not present in a number of first and second generation clocks (including Horvath’s and Hannum’s clock and PhenoAge) that were measured in the same samples [47].

Taken together, the first-generation clocks, i.e. the 7-CpG clock, Horvath’s clock and Hannum’s clock, do not seem to represent biological age very well in the context of the functional domains of aging analyzed here. Whether this is also true for second-generation clocks needs to be assessed in future studies. Marioni and colleagues proposed the following possible explanations for the lack of associations between DNAmAA and markers of physical and mental fitness: (i) such a (longitudinal) association does not exist, (ii) the change in the analyzed variables over time is too small and (iii) the analyzed cohort is too healthy [19]. All three arguments also apply to our study as well. With respect to the latter argument, for example, 28.5% of participants at baseline (mean age 68.3 years) and 53.5% of participants at follow-up (mean age 75.6 years) are classified as frail/pre-frail according to the Fried criteria. This outcome is (well) below the average that was reported in comparable cohorts of similar mean chronological age before. For instance, Ahrenfeldt and colleagues reported 54.6% of the 113,299 participants of the SHARE study (244,258 observations, 45.7% female, mean age: 66.2 years) to be pre-frail or frail according to the Fried criteria [50]. Likewise, Fried and colleagues themselves [32] found 48% of 3,599 participants between 65 and 74 years to be frail or pre-frail.

However, the lack of observable associations between molecular biomarkers and mental and physical capacity as assessed here does not generally rule out that such an association exists and may also depend on limitations of the study design. For this study, we would like to emphasize the following limitations. First, all of our (negative) conclusions from logistic regression analyses are based on the cut-off values chosen for the various domains. We note, however, that cut-offs used in this study are all well established and evaluated and continuous values were analyzed as well. Still, the assessments employed here represent only one possible means of quantifying very complex clinical and functional aspects and are only weakly correlated among themselves. Second, another limitation of our study relates to the small age range covered by the participants. It is possible that the analyzed biomarkers of aging only reveal their potential association with the analyzed domains of aging in an older cohort or a cohort with an on average higher degree of impairment. Therefore, it would be of interest to repeat these analyses in studies with participants showing a higher morbidity burden. Third, the lack to detect significant associations could simply reflect a lack in power. While formal power analyses are difficult to conduct in the setting of multiple variable analysis with a number of unproven and unknown assumptions, we note that the sample size analyzed here is in the middle range among datasets with a similar breadth of both DNA methylation and clinical data available (overview on previous studies in Supplementary Table 8). Notwithstanding, limited power will likely have affected some of our conclusions. Fourth, our analyses could be biased by not considering all covariates relevant in this context. However, this is true for nearly every study using DNA methylation data as molecular marker. We note that we went to great lengths to review the current literature and choose the most meaningful covariates of likely relevance in this setting that were available in our dataset, so the impact of this limitation should be minimized. In this context we would also like to note that the understanding of the process that underlies age-dependent epigenetic changes, e.g. at CpG sites considered for epigenetic clock calculations, is currently still very limited. Therefore, explanations for the observed lack of association between the biomarkers and results in functional assessments must remain speculative. In any case, future work needs to re-assess this question in independent and sufficiently sized longitudinal cohorts to evaluate the predictive value of epigenetic age as a biomarker in this context.

Strengths of our study include the availability of DNAm data generated by two different experimental methods. In case of the 7-CpG clock, we also had DNAm measurements at two time-points enabling us to perform longitudinal analyses, which, to our knowledge, were not investigated by any other study before. Third, the BASE-II/GendAge-cohort is a carefully ascertained and thoroughly phenotyped dataset which provides detailed information on several distinct domains of mental and physical health in aging individuals, allowing us to assess functional capacity comprehensively.

## Conclusion

None of our longitudinal functional capacity assessments were associated with rLTL or 7-CpG DNAmAA at baseline. Furthermore, cross-sectional analyses also showed no association between the same variables and DNAmAA derived from five different epigenetic age estimates. The only possible exception was a weak association between frailty and GrimAge DNAmAA, which needs to be assessed in future work. Thus, overall, DNAmAA does not appear to be a good biomarker of age-associated decline in physical or mental capacity, at least not in the group of comparably healthy older adults investigated here. Additional longitudinal studies with a higher morbidity burden and larger sample size are needed to further evaluate the utility of DNAm age as a molecular biomarker of longitudinal trajectories of aging-related phenotypes.

## Data Availability

Data are available upon reasonable request. Interested investigators are invited to contact the study coordinating PI Ilja Demuth at to obtain additional information about the GendAge study and the data-sharing application form.

## Acknowledgments

-

## Author contributions

Conceived and designed the study: VMV and ID. Contributed study specific data: all authors. Analyzed the data: VMV and YS. Wrote the manuscript: VMV and ID. All authors revised and approved the manuscript.

