## Supplementary figures and images for "Relationship between five Epigenetic Clocks, Telomere Length and Functional Capacity assessed in Older Adults: Cross-sectional and Longitudinal Analyses"

### Supplementary Figure 1

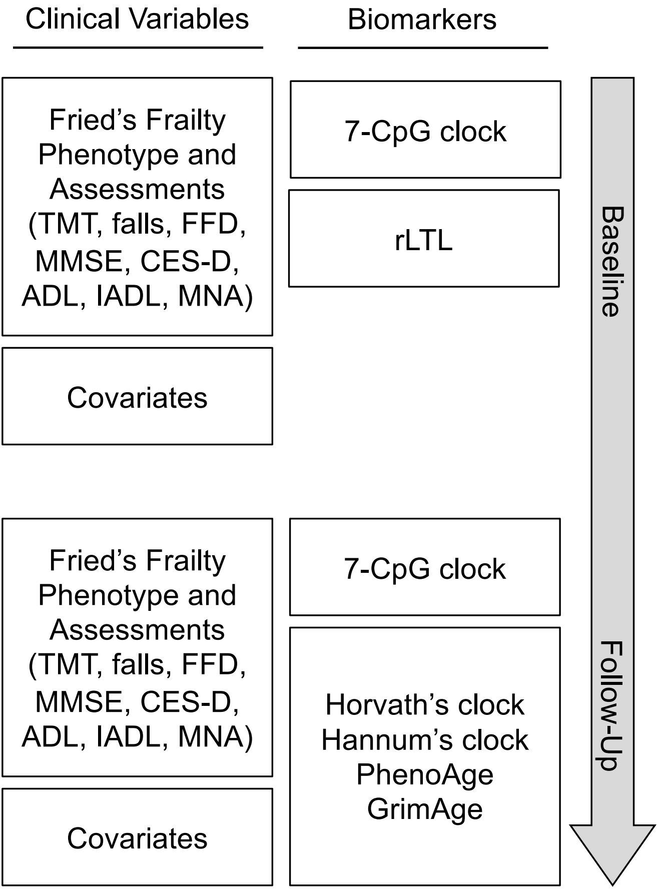
