## Supplementary Table 1 for "Relationship between five Epigenetic Clocks, Telomere Length and Functional Capacity assessed in Older Adults: Cross-sectional and Longitudinal Analyses"

|  | Women and Men | | | | |  | Women | | | | |  | Men | | | | |  |  |
| --- | --- | --- | --- | --- | --- | --- | --- | --- | --- | --- | --- | --- | --- | --- | --- | --- | --- | --- | --- |
|  | n | Mean | SD | Min | Max |  | n | Mean | SD | Min | Max |  | n | Mean | SD | Min | Max |  | p-value |
| Horvath DNAm age | 1088 | 72.44 | 4.64 | 59.86 | 100.41 |  | 561 | 71.94 | 4.45 | 59.86 | 100.41 |  | 511 | 73.06 | 4.79 | 61.43 | 95.14 |  | <0.001 |
| Hannum DNAm age | 1088 | 63.22 | 4.85 | 48.48 | 97.83 |  | 561 | 62.45 | 4.56 | 50.72 | 82.79 |  | 511 | 64.09 | 4.99 | 51.44 | 97.83 |  | <0.001 |
| PhenoAge DNAm age | 1088 | 61.24 | 6.34 | 42.18 | 93.38 |  | 561 | 60.56 | 6.20 | 43.75 | 93.38 |  | 511 | 61.95 | 6.41 | 42.18 | 84.04 |  | <0.001 |
| GrimAge DNAm age | 1088 | 75.61 | 4.53 | 61.86 | 93.13 |  | 561 | 74.17 | 3.99 | 63.15 | 89.93 |  | 511 | 77.23 | 4.53 | 63.98 | 93.13 |  | <0.001 |
| Horvath DNAmAA | 1067 | 0.03 | 4.04 | -12.31 | 23.45 |  | 549 | -0.35 | 3.96 | -12.31 | 23.45 |  | 502 | 0.53 | 4.06 | -8.94 | 17.44 |  | <0.001 |
| Hannum DNAmAA | 1067 | 0.01 | 3.89 | -10.80 | 28.57 |  | 549 | -0.69 | 3.66 | -10.80 | 12.73 |  | 502 | 0.79 | 3.96 | -9.32 | 28.57 |  | <0.001 |
| PhenoAge DNAmAA | 1067 | 0.04 | 5.42 | -16.54 | 25.80 |  | 549 | -0.50 | 5.40 | -16.54 | 25.80 |  | 502 | 0.59 | 5.39 | -13.51 | 20.94 |  | 0.001 |
| GrimAge DNAmAA | 1067 | 0.03 | 3.39 | -10.83 | 12.85 |  | 549 | -1.28 | 2.94 | -10.83 | 10.71 |  | 502 | 1.48 | 3.27 | -8.17 | 12.85 |  | <0.001 |

**Supplementary Table 1: Descriptive Statistics of the four epigenetic age estimates derived from Illumina methylation data.**

Note: DNAm age: DNA methylation age; DNAmAA: DNA methylation age acceleration.
