## Supplementary Table 2 for "Relationship between five Epigenetic Clocks, Telomere Length and Functional Capacity assessed in Older Adults: Cross-sectional and Longitudinal Analyses"

**Supplementary Table 2: Descriptive statistics showing the number of participants at follow-up who show impairment only in a singular assessment (“isolated impairment”) and their proportion of all observed participants and of all participants showing impairment.**

|  | n  (isolated impairment) | n  (not-impaired participants) | n  (impaired participants) | n  (all participants) | n (isolated impairment)/  n (all observations) | n (isolated impairment)/  n (impaired participants) |
| --- | --- | --- | --- | --- | --- | --- |
| Frailty score | 85 | 502 | 568 | 1070 | 7.9% | 15.0% |
| Tinetti mobility test | 3 | 968 | 83 | 1051 | 0.3% | 3.6% |
| Falls in past 12 months | 21 | 631 | 237 | 868 | 2.4% | 8.9% |
| Finger-floor distance | 182 | 404 | 664 | 1068 | 17.0% | 27.4% |
| MMSE | 1 | 1052 | 19 | 1071 | 0.1% | 5.3% |
| CES-D | 25 | 818 | 254 | 1072 | 2.3% | 9.8% |
| ADL | 3 | 1028 | 49 | 1077 | 0.3% | 6.1% |
| IADL | 0 | 1070 | 7 | 1077 | 0.0% | 0.0% |
| MNA | 14 | 932 | 117 | 1049 | 1.3% | 12.0% |

Note: MMSE: Mini-Mental State Examination, CES-D: Center for Epidemiologic Studies Depression Scale, ADL: Activities of Daily Living, IADL: Instrumented ADL, MNA: Mini Nutritional Assessment.
