## Supplementary Table 3 for "Relationship between five Epigenetic Clocks, Telomere Length and Functional Capacity assessed in Older Adults: Cross-sectional and Longitudinal Analyses"

**Supplementary Table 3: Number of participants showing impairment in zero to seven assessments at baseline and at follow-up. The difference between the percentage of all participants at baseline and at follow-up is given in percentage points in the last column.**

|  | Baseline | | | | | | | | | | | |  | | Follow-up | | | | | | | | | | | | |  | | | |
| --- | --- | --- | --- | --- | --- | --- | --- | --- | --- | --- | --- | --- | --- | --- | --- | --- | --- | --- | --- | --- | --- | --- | --- | --- | --- | --- | --- | --- | --- | --- | --- |
|  | Women and Men | |  | Women | |  | | Men | | |  | Women and Men | | | | | |  | | Women | | |  | | Men | | | |  | | |
| Number of assessments  showing impairment | n | % |  | n | % | |  | | n | % | | | |  | | n | % | |  | | n | % | |  | | n | % | | |  | Difference  (percentage points) |
| 0 | 181 | 16.71 |  | 126 | 22.38 | |  | | 55 | 10.58 | | | |  | | 131 | 12.10 | |  | | 88 | 15.63 | |  | | 43 | 8.27 | | |  | -4.62 |
| 1 | 433 | 39.98 |  | 203 | 36.06 | |  | | 230 | 44.23 | | | |  | | 334 | 30.84 | |  | | 173 | 30.73 | |  | | 161 | 30.96 | | |  | -9.14 |
| 2 | 306 | 28.25 |  | 152 | 27.00 | |  | | 154 | 29.62 | | | |  | | 327 | 30.19 | |  | | 149 | 26.47 | |  | | 178 | 34.23 | | |  | 1.94 |
| 3 | 127 | 11.73 |  | 63 | 11.19 | |  | | 64 | 12.31 | | | |  | | 188 | 17.36 | |  | | 91 | 16.16 | |  | | 97 | 18.65 | | |  | 5.63 |
| 4 | 31 | 2.86 |  | 16 | 2.84 | |  | | 15 | 2.88 | | | |  | | 76 | 7.02 | |  | | 41 | 7.28 | |  | | 35 | 6.73 | | |  | 4.16 |
| 5 | 3 | 0.28 |  | 1 | 0.18 | |  | | 2 | 0.38 | | | |  | | 21 | 1.94 | |  | | 17 | 3.02 | |  | | 4 | 0.77 | | |  | 1.66 |
| 6 | 1 | 0.09 |  | 1 | 0.18 | |  | | 0 | 0.00 | | | |  | | 5 | 0.46 | |  | | 3 | 0.53 | |  | | 2 | 0.38 | | |  | 0.37 |
| 7 | 1 | 0.09 |  | 1 | 0.18 | |  | | 0 | 0.00 | | | |  | | 1 | 0.09 | |  | | 1 | 0.18 | |  | | 0 | 0.00 | | |  | 0.00 |
| n (total) | 1083 |  |  | 563 |  | |  | | 520 |  | | | |  | | 1083 |  | |  | | 563 |  | |  | | 520 |  | | |  |  |
| Average impairments  per person | 1.46 |  |  | 1.38 |  | |  | | 1.54 |  | | | |  | | 1.84 |  | |  | | 1.81 |  | |  | | 1.88 |  | | |  |  |
