## Supplementary Table 4 for "Relationship between five Epigenetic Clocks, Telomere Length and Functional Capacity assessed in Older Adults: Cross-sectional and Longitudinal Analyses"

**Supplementary Table 4: Individual logistic regression analyses of functional assessments at follow-up on rLTL and DNAmAA and adjusted for genetic PCs, sex (if applicable), alcohol consumption, smoking behavior, morbidity index and body mass index.** **Biomarkers of aging and all covariates were measured at baseline.**

|  | Women and Men | | | | |  | Women | | | | |  | Men | | | | |
| --- | --- | --- | --- | --- | --- | --- | --- | --- | --- | --- | --- | --- | --- | --- | --- | --- | --- |
| Assessments | Estimate | SE | OR | p | n |  | Estimate | SE | OR | p | n |  | Estimate | SE | OR | p | n |
|  | rLTL | | | | | | | | | | | | | | | | |
| Frailty score | 0.19 | 0.34 | 1.21 | 0.58 | 749 |  | -0.08 | 0.56 | 0.93 | 0.89 | 381 |  | 0.33 | 0.43 | 1.39 | 0.44 | 368 |
| Tinetti mobility test | -0.04 | 0.68 | 0.96 | 0.95 | 743 |  | 0.39 | 1.05 | 1.47 | 0.71 | 381 |  | -0.51 | 0.94 | 0.60 | 0.59 | 362 |
| Falls in past 12 months | 0.01 | 0.41 | 1.01 | 0.99 | 608 |  | -0.05 | 0.63 | 0.95 | 0.93 | 327 |  | 0.13 | 0.55 | 1.14 | 0.82 | 281 |
| Finger-floor distance | -0.23 | 0.38 | 0.79 | 0.54 | 749 |  | -0.11 | 0.54 | 0.90 | 0.84 | 381 |  | -0.33 | 0.54 | 0.72 | 0.55 | 368 |
| MMSE | -0.40 | 1.23 | 0.67 | 0.75 | 750 |  | 2.51 | 2.01 | 12.29 | 0.21 | 383 |  | -2.86 | 2.04 | 0.06 | 0.16 | 367 |
| CES-D | 0.20 | 0.40 | 1.23 | 0.61 | 751 |  | -0.49 | 0.61 | 0.61 | 0.42 | 382 |  | 0.75 | 0.55 | 2.13 | 0.17 | 369 |
| ADL | -0.89 | 0.83 | 0.41 | 0.29 | 754 |  | 0.33 | 1.24 | 1.39 | 0.79 | 384 |  | -2.11 | 1.23 | 0.12 | 0.09 | 370 |
| MNA | -0.45 | 0.55 | 0.64 | 0.42 | 738 |  | 0.01 | 0.83 | 1.01 | 0.99 | 374 |  | -0.99 | 0.77 | 0.37 | 0.20 | 364 |
|  | 7-CpG DNAm Age Acceleration | | | | | | | | | | | | | | | | |
| Frailty score | 0.00 | 0.01 | 1.00 | 0.80 | 713 |  | -0.03 | 0.02 | 0.97 | 0.05 | 363 |  | 0.02 | 0.02 | 1.03 | 0.13 | 350 |
| Tinetti mobility test | 0.03 | 0.02 | 1.03 | 0.27 | 708 |  | 0.02 | 0.03 | 1.02 | 0.61 | 363 |  | 0.03 | 0.03 | 1.03 | 0.35 | 345 |
| Falls in past 12 months | 0.01 | 0.01 | 1.01 | 0.68 | 582 |  | 0.01 | 0.02 | 1.01 | 0.65 | 313 |  | 0.00 | 0.02 | 1.00 | 0.90 | 269 |
| Finger-floor distance | 0.00 | 0.01 | 1.00 | 0.79 | 714 |  | 0.00 | 0.02 | 1.00 | 0.87 | 364 |  | 0.02 | 0.02 | 1.02 | 0.43 | 350 |
| MMSE | -0.03 | 0.04 | 0.97 | 0.53 | 714 |  | 0.03 | 0.07 | 1.03 | 0.70 | 365 |  | -0.09 | 0.06 | 0.91 | 0.14 | 349 |
| CES-D | -0.02 | 0.01 | 0.98 | 0.11 | 715 |  | -0.02 | 0.02 | 0.98 | 0.27 | 364 |  | -0.02 | 0.02 | 0.98 | 0.26 | 351 |
| ADL | 0.00 | 0.03 | 1.00 | 0.94 | 718 |  | -0.07 | 0.04 | 0.93 | 0.09 | 366 |  | 0.08 | 0.04 | 1.08 | 0.08 | 352 |
| MNA | 0.00 | 0.02 | 1.00 | 0.94 | 703 |  | 0.01 | 0.03 | 1.01 | 0.69 | 356 |  | 0.00 | 0.03 | 1.00 | 0.94 | 347 |

Note: MMSE: Mini-Mental State Examination, CES-D: Center for Epidemiologic Studies Depression Scale, ADL: Activities of Daily Living, MNA: Mini Nutritional Assessment.
