## Supplementary Table 5 for "Relationship between five Epigenetic Clocks, Telomere Length and Functional Capacity assessed in Older Adults: Cross-sectional and Longitudinal Analyses"

**Supplementary Table 5: Sex-stratified individual multiple linear regression analyses of functional assessments at follow-up on rLTL and DNAmAA and adjusted for genetic PCs, alcohol consumption, smoking behavior (packyears), morbidity index and body mass index. Biomarkers of aging and all covariates were measured at baseline.**

|  |  | Women | | | |  | Men | | | |
| --- | --- | --- | --- | --- | --- | --- | --- | --- | --- | --- |
| Assessments |  | β | SE | p-value | n |  | β | SE | p-value | n |
| rLTL | | | | | | | | | | |
| Frailty score |  | -0.13 | 0.62 | 0.84 | 381 |  | 0.16 | 0.17 | 0.35 | 368 |
| Tinetti mobility test |  | 1.91 | 3.08 | 0.54 | 381 |  | 0.16 | 0.50 | 0.75 | 362 |
| Finger-floor distance |  | -0.30 | 0.40 | 0.44 | 383 |  | -3.12 | 2.58 | 0.23 | 368 |
| MMSE |  | 0.44 | 1.04 | 0.67 | 382 |  | 0.31 | 0.34 | 0.37 | 367 |
| CES-D |  | -0.36 | 0.82 | 0.66 | 384 |  | 0.94 | 0.70 | 0.18 | 369 |
| ADL |  | 0.00 | 0.07 | 0.98 | 382 |  | 0.26 | 1.09 | 0.81 | 370 |
| MNA |  | -0.01 | 0.01 | 0.07 | 363 |  | 0.11 | 0.42 | 0.80 | 364 |
| 7-CpG DNAm Age Acceleration | | | | | | | | | | |
| Frailty score |  | -0.01 | 0.02 | 0.68 | 363 |  | 0.01 | 0.01 | 0.24 | 350 |
| Tinetti mobility test |  | -0.10 | 0.09 | 0.29 | 364 |  | -0.02 | 0.02 | 0.29 | 345 |
| Finger-floor distance |  | 0.00 | 0.01 | 0.83 | 365 |  | 0.03 | 0.10 | 0.72 | 350 |
| MMSE |  | -0.02 | 0.03 | 0.44 | 364 |  | 0.01 | 0.01 | 0.49 | 349 |
| CES-D |  | 0.02 | 0.02 | 0.35 | 366 |  | -0.01 | 0.03 | 0.71 | 351 |
| ADL |  | 0.00 | 0.00 | 0.40 | 364 |  | -0.10 | 0.04 | 0.02 | 352 |
| MNA |  | 0.00 | 0.02 | 0.91 | 356 |  | 0.00 | 0.02 | 0.76 | 347 |

Note: MMSE: Mini-Mental State Examination, CES-D: Center for Epidemiologic Studies Depression Scale, ADL: Activities of Daily Living, MNA: Mini Nutritional Assessment.
