## Supplementary Table 7 for "Relationship between five Epigenetic Clocks, Telomere Length and Functional Capacity assessed in Older Adults: Cross-sectional and Longitudinal Analyses"

**Supplementary Table 7: Individual multiple linear regression analyses of functional assessments on DNAmAA of five different epigenetic clocks and genetic PCs, alcohol consumption, smoking behavior, morbidity index and body mass index in the all-male subgroup.**

| Independent Variable | Dependent Variable | Estimate | SE | p-value |  | n |
| --- | --- | --- | --- | --- | --- | --- |
| 7-CpG IEAA | Frailty score | 0.01 | 0.01 | 0.16 |  | 373 |
|  | Tinetti mobility test | 0.00 | 0.02 | 0.85 |  | 364 |
|  | Finger-floor distance | 0.00 | 0.10 | 0.96 |  | 373 |
|  | MMSE | 0.02 | 0.01 | 0.20 |  | 370 |
|  | CES-D | 0.03 | 0.03 | 0.36 |  | 375 |
|  | ADL | -0.07 | 0.04 | 0.09 |  | 373 |
|  | MNA | -0.01 | 0.02 | 0.74 |  | 365 |
| Horvath's IEAA | Frailty score | 0.00 | 0.01 | 0.88 |  | 371 |
|  | Tinetti mobility test | 0.03 | 0.02 | 0.31 |  | 362 |
|  | Finger-floor distance | -0.20 | 0.16 | 0.20 |  | 371 |
|  | MMSE | 0.01 | 0.02 | 0.67 |  | 367 |
|  | CES-D | -0.02 | 0.04 | 0.61 |  | 373 |
|  | ADL | 0.07 | 0.06 | 0.28 |  | 371 |
|  | MNA | 0.00 | 0.02 | 0.99 |  | 363 |
| Hannum's IEAA | Frailty score | 0.00 | 0.01 | 0.69 |  | 371 |
|  | Tinetti mobility test | -0.01 | 0.03 | 0.63 |  | 362 |
|  | Finger-floor distance | -0.24 | 0.16 | 0.14 |  | 371 |
|  | MMSE | -0.02 | 0.02 | 0.32 |  | 367 |
|  | CES-D | 0.00 | 0.05 | 0.95 |  | 373 |
|  | ADL | -0.01 | 0.07 | 0.86 |  | 371 |
|  | MNA | -0.01 | 0.02 | 0.57 |  | 363 |
| PhenoAge IEAA | Frailty score | 0.00 | 0.01 | 0.73 |  | 371 |
|  | Tinetti mobility test | 0.02 | 0.02 | 0.39 |  | 362 |
|  | Finger-floor distance | 0.00 | 0.12 | 1.00 |  | 371 |
|  | MMSE | 0.01 | 0.02 | 0.59 |  | 367 |
|  | CES-D | -0.02 | 0.03 | 0.66 |  | 373 |
|  | ADL | -0.03 | 0.05 | 0.51 |  | 371 |
|  | MNA | 0.00 | 0.02 | 0.86 |  | 363 |
| GrimAge IEAA | Frailty score | 0.02 | 0.01 | 0.18 |  | 371 |
|  | Tinetti mobility test | -0.04 | 0.03 | 0.30 |  | 362 |
|  | Finger-floor distance | -0.16 | 0.21 | 0.46 |  | 371 |
|  | MMSE | 0.01 | 0.03 | 0.75 |  | 367 |
|  | CES-D | 0.06 | 0.06 | 0.34 |  | 373 |
|  | ADL | -0.05 | 0.09 | 0.57 |  | 371 |
|  | MNA | -0.06 | 0.03 | 0.09 |  | 363 |
