## Supplementary Table 8 for "Relationship between five Epigenetic Clocks, Telomere Length and Functional Capacity assessed in Older Adults: Cross-sectional and Longitudinal Analyses"

**Supplementary Table 8: Overview on previous studies that analyzed the cross-sectional association between epigenetic clocks and physical and cognitive capacity.**

| Study | N | Age | Female | Covariates | Clocks | Assessments | Key Results |
| --- | --- | --- | --- | --- | --- | --- | --- |
| Current Study | N=1100 | Age range: 65-94 years, 75.6 (3.8) years mean age (SD) | 52.1% | Genetic PCs, sex, alcohol consumption, smoking behavior, morbidity index, body mass index (BMI) | 7-CpG clock (IEAA),  Horvath’s clock (IEAA),  Hannum’s clock (IEAA),  GrimAge (IEAA),  PhenoAge (IEAA) | Fried’s frailty score, TMT, falls in the past 12 months (yes/no), Finger-floor distance, MMSE, CES-D, ADL, IADL, MNA | No association |
| Vershoor et al. [1] | N=1446 | mean age (SD): 63 (10.3) years | 50.6% | Chronological age, sex, education, income, smoking, physical activity, diet | Horvath’s clock (DNAmAA),  Hannum’s clock (DNAmAA),  PhenoAge (DNAmAA),  GrimAge (DNAmAA),  Lin’s clock (DNAmAA),  Yang’s clock (DNAmAA),  Dunedin PoAm (DNAmAA),  Zhang’s clock (DNAmAA) | 76 -item Frailty Index | Cross-sectional association with GrimAge |
| McCrory et al. [2] | N=490 | mean age (SD): 62.3 (8.3) years | 50.2% | Chronological age, sex, white blood cell counts, height (for walking speed and grip strength), life course social class trajectory, smoking, physical activity, body mass index (BMI) | Horvath’s clock (IEAA),  Hannum’s clock (IEAA),  PhenoAge (IEAA),  GrimAge (IEAA) | Walking speed, grip strength, Fried frailty score, MMSE, MOCA, Sustained Attention Reaction Time, 2-choice reaction time | Association between GrimAge and Fried’s frailty score after adjustment for covariates |
| Maddock et al. [3] | NSHD 53y: n=1375  NSHD 60-64y: n=672  NCDS: n=240  Twins UK: n=120 | 53 years  60-64 years  45 years  mean age (SD): 64.6 (9.3) years | 52.4%  48.6%  53.3%  100% | Body mass index (BMI), height, smoking status, socioeconomic position (occupational social class or income) | Horvath’s clock (DNAmAA),  Hannum’s clock (DNAmAA),  PhenoAge (DNAmAA),  GrimAge (DNAmAA) | Grip strength, chair rise speed, episodic memory, mental speed | Associations between PhenoAge and chair rise speed and between GrimAge and cognitive capacity measures |
| Gale et al. [4] | N=791 | 70 years | 50.3% | Chronological age, sex, smoking, alcohol intake, chronic disease | Horvath’s clock (IEAA)  Hannum’s clock (EEAA) | Fried’s frailty score | Association with Hannum’s EEAA |
| Kim et al. [5] | N=262 | Age range: 60 – 103 years,  mean age (SD): 86 (10) years | 60.7% | Chronological age | Horvath’s clock (DNAm age) | 34-items Frailty Index | No association after adjustment for age |
| Breitling et al. [6] | N=1820 (dataset 1: n=969, dataset 2: n=851) | 50-75 years, dataset 1 mean age(SD): 62.1 (6.5) years,  Dataset 2 mean age (SD): 63.0 (6.7) years | Dataset 1: 50.0%,  Dataset 2: 54.5% | Chronological age, sex, leukocyte cell distribution, smoking, alcohol intake, history of cancer | Horvath’s clock (difference based DNAmAA) | 34-items Frailty Index | Increase of one point in Frailty Index for every 11.6 years of DNAmAA |

Note: MOCA: Montreal Cognitive Assessment, MMSE: Mini-Mental State Examination, CES-D: Center for Epidemiologic Studies Depression Scale, ADL: Activities of Daily Living, IADL: Instrumented ADL, MNA: Mini Nutritional Assessment, IEAA: intrinsic epigenetic age acceleration, DNAm age: DNA methylation age, DNAmAA: DNA methylation age acceleration.
